# Development and Pilot Testing for a Novel Shared Decision-Making Tool for Tracheostomy Decision-Making

**DOI:** 10.1101/2025.02.24.25322755

**Authors:** Anuj B Mehta, Steven Lockhart, Ivor S Douglas, Meredith Mealer, Daniel D Matlock

**Author notes:** **Corresponding Author**: Anuj B Mehta, MD. 777 Bannock St. E320. Denver, CO, 80204. **Funding:** ABM is supported by NIH K23HL141704 (Primary funding source). **Conflict of Interest Declaration**: All authors attest to the fact that they had no financial or intellectual conflicts of interest. **Author Contributions**: ABM, ISD, and DDM conceived the study. ABM and SL were responsible for data collection and analysis. ABM and DDM were responsible for quantitative data interpretation. ABM, SL, and MM were responsible for qualitative data interpretation. ABM drafted the article. All authors provided critical revisions and meaningful input for the final draft. ABM had full access to all the data in the study and takes responsibility for the integrity of the data and the accuracy of the data analysis. All authors approved the final draft of the manuscript.

## Abstract

**Background:** Tracheostomy and prolonged mechanical ventilation decision-making is one of the most emotionally difficult decisions facing surrogate decision-makers in health care. Often, surrogates face decisions between the potential for prolonged life support verses transitions to comfort measures and possible death. Despite more than two decades of research, major gaps exist in improving the decision-making process.

**Objective:** Develop and pilot testing a novel shared decision-making tool for tracheostomy and prolonged mechanical ventilation.

**Methods:** Development of the novel web-based conversation tool called TRACH-Support was an iterative process engaging key stakeholders (patients, surrogates, critical care providers, and shared decision-making experts) at multiple points. Development of the website used a Human-Centered Design approach with modern graphics and website interfaces. Pragmatic pilot testing was a mixed methods approach recruiting surrogates, providers, nurses, and respiratory therapists. Primary quantitative outcomes included Usability (System Usability Scale (SUS)) and Acceptability (Acceptability of Intervention (AIM)) measures with multiple secondary outcomes. Qualitative interviews used a Think Aloud approach and matrix analysis methodology.

**Results:** A total of 86 participants were recruited for the quantitative survey with 10 surrogates and 10 providers completing qualitative interviews. Mean SUS score among all participants was 68.2/100 (SD=10.7) but surrogates specifically had a mean SUS=74.2/100 (74.2). The overall mean AIM score was 4.2/5 (SD=0.8) and 79.4% of all participants viewed TRACH-Support as “Acceptable” or “Very Acceptable”. Qualitative interviews indicated that TRACH-Support had high Usability and Acceptability. Customizability, pictures, novel outcomes, and the organization were all features that contributed to participant views. Participants also suggested several modifications including reducing the word count, adding video testimonials, and adding information on how faith/religion may play a role in decision-making for some.

**Conclusions:** TRACH-Support, a novel, web-based, customizable and personalizable conversation tool for tracheostomy and prolonged mechanical ventilation was developed according to the most rigorous standards for decision-support tools. It had high Usability and Acceptability as assessed by quantitative and qualitative measures. Future large-scale testing is needed to assess real-world effectiveness and implementation.

## Introduction

Annually, more than 100,000 adults in the United States receive a tracheostomy to facilitate prolonged mechanical ventilation (PMV).(1–3) Tracheostomy can be a life-prolonging intervention for patients who need PMV as prolonged endotracheal intubation carries significant risk. Some reports suggest that patients who survive tracheostomy to the point of being able to return home have a positive view of their experience.(4) However, multiple studies also suggest that patients who receive a tracheostomy have high short- and long-term mortality and that while alive, many suffer significant cognitive and functional disabilities with repeated hospitalizations.(5–11) When asked, many patients view the thought of being attached to machines as being “worse than death”.(12)

Tracheostomy represents a branch point in the care of patients who require prolonged mechanical ventilation. Most often, the choice rests between transitions to palliative options verses ongoing artificial life support. Unlike many other decisions in health care, tracheostomy decisions almost universally fall to surrogate decision-makers as the patient is intubated and most often sedated. There is a large body of evidence that suggests decision-making for critically ill patients is less than ideal and that patients and families are, at times, deeply dissatisfied with the communication and information they receive.(13–16) One specific study among surrogates making tracheostomy decisions highlighted core gaps in the decision-making process.(17) The current state of decision-making is inadequate to meet the needs of most surrogates and raises the real risk of care that is not goal-concordant.

However, decision-making for critically ill patients is a two-way street with key involvement from providers. Providers have themselves identified gaps in their ability to guide surrogates during this process and many providers have indicated that their struggles in these areas contribute to burnout.(17,18) It is also possible that the lack of a roadmap to help providers with the decision-making process may contribute to the wide variation seen in tracheostomy practices across different hospitals.(19–23) A previous attempt to improve the decision-making process utilizing a decision-aid did not show significant changes in decisional conflict but its foundational development was limited and did not incorporate more recent understandings of time trials in critical care.(24–29)

Core improvements in tracheostomy decision-making are necessary to meet the needs of patients, surrogates, providers, and the wider health care team and to ensure goal-concordant care. Herein, we describe the development and pilot testing for TRACH-Support, a novel web-based personalized conversation tool to support team-based shared decision-making for tracheostomy and PMV.

## Methods

### TRACH-Support Development

The development was governed by the International Patient Decision-Aid Standards (IPDAS) and the Ottawa Decision Support Framework (ODSF).(30–34) The core decision addressed by TRACH-Support was tracheostomy/PMV but unlike prior decision support tools, the options proposed incorporated a broader understanding of prolonged intubation, the key introduction of time trials associated with tracheostomy as recommended by professional societies, and comfort focused approaches (**Figure 1**).(26–29,35) An iterative process engaging key stakeholders at multiple points was employed to arrive at the final version of TRACH-Support used for pilot testing (**Figure 2**). Key domains to be addressed by TRACH-Support were based on our prior qualitative decisional needs assessment (**Table 1**).(17) From the outset, the plan was to develop an online, interactive, and personalizable tool that addressed previous limitations of decision-support tools.

**Figure 1:**
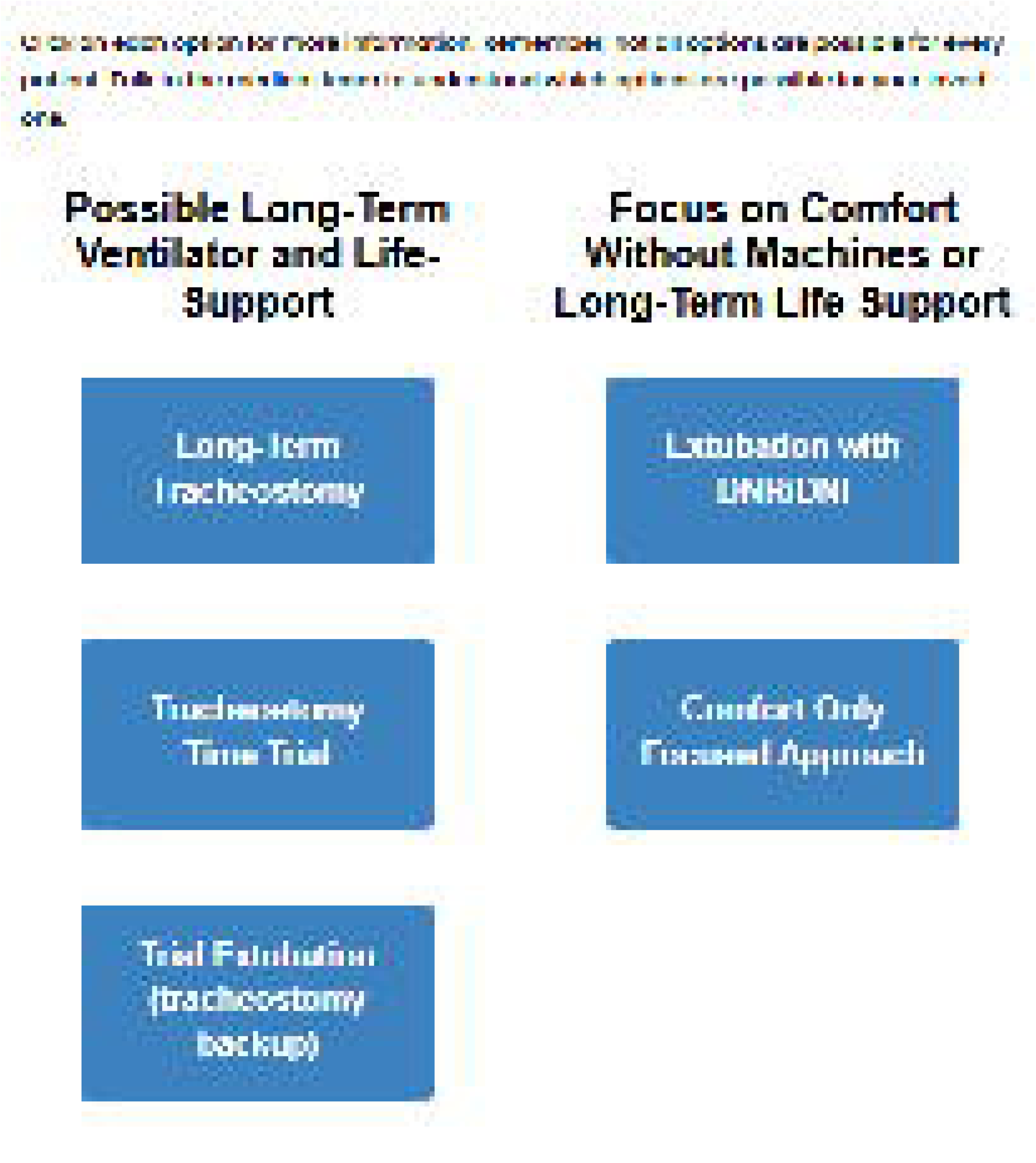
TRACH-Support Options Matrix. – TRACH-Support focuses on options surrounding tracheostomy and prolonged mechanical ventilation. The options sit along a values spectrum ranging from the possibility of long-term life support at one end and a focus on comfort at the other end. The options within the scope of possible long-term life support include tracheostomy with long-term ventilation, the novel idea of a tracheostomy time trial, and a trial extubation but with the option of reintubation and tracheostomy. On the side of focusing on comfort two options are presented both of which include extubation. The first introduces the idea of extubation with a Do-Not-Resuscitate/Do-Not-Intubate order but with instructions to continue to other medical treatments (e.g., antibiotics, vasopressors, etc.). The second option includes a full transition to comfort focused measures only with a palliative extubation. This matrix moves beyond the previous dichotomous decision options of tracheostomy verses comfort measures only. Abbreviations: DNR/DNI - Do-Not-Resuscitate/Do-Not-Intubate

**Figure 2:**
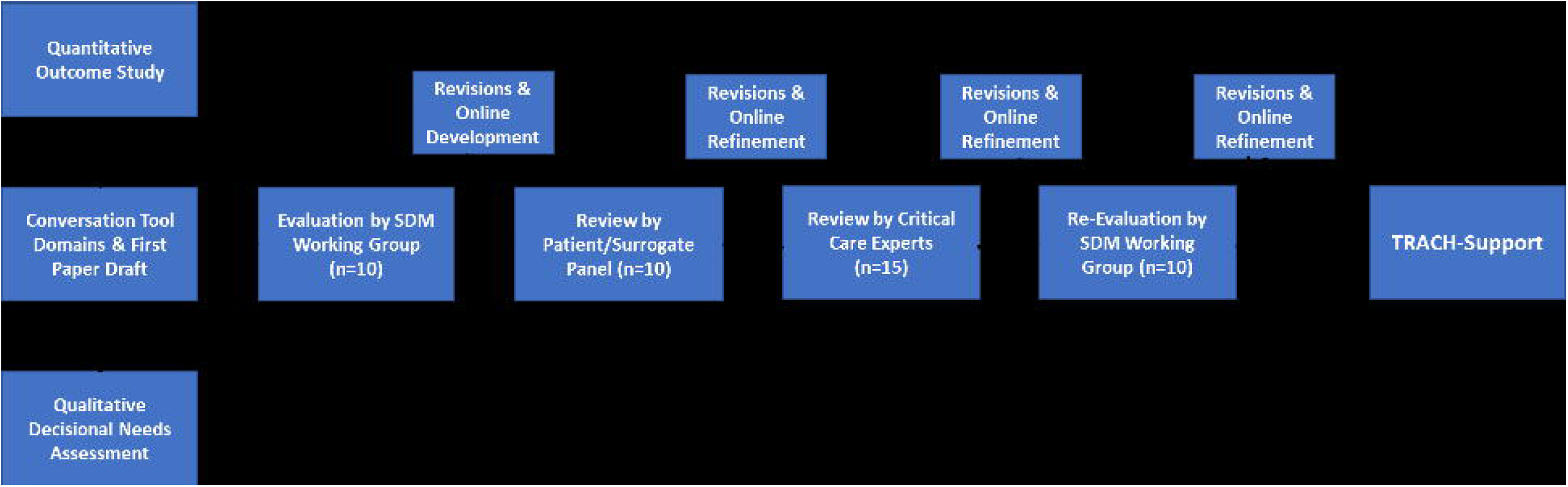
Iterative Development of TRACH-Support. – TRACH-Support was based on foundational quantitative and qualitative studies. The development was iterative with engagement from multiple stakeholders including shared decision-making experts, patients, surrogates, and critical care experts. Abbreviations: SDM – shared decision-making.

**Table 1:** Key Domains to be Addressed by TRACH-Support Identified From Prior Work.

| **Domain** | **Description** |
| --- | --- |
| **Nature of the decision** | Description of the limitations of endotracheal intubation, options around prolonged mechanical ventilation including tracheostomy, and exploration of comfort-focused options |
| **Clarification about role of surrogate decision-makers** | In preliminary work, many surrogate decision-makers expressed confusion about their role |
| **Patient values and preferences** | In some cases, surrogates know what a patient would want. However, in most cases, such discussions have never happened and surrogates wished for a guide on how to infer a patient's wishes. Providers expressed a strong-desire for a roadmap to help surrogates identify likely values and goals |
| **Risks and benefits** | Many surrogates indicated that there was insufficient information about potential risks and benefits |
| **Long-term implications** | Clarification of the long-term implications of tracheostomy including the potential for recovery vs potential for prolonged hospitalization. Within this domain, surrogates wanted information about long-term hospitalization options and considerations. |
| **Outcomes** | Both surrogates and providers indicated that there was limited information about outcomes after a tracheostomy |
| **Images** | Most providers indicated they had never shown surrogates a picture of someone with a tracheostomy. Surrogates also indicated that pictures of tracheostomies would be extremely beneficial. |

### Knowledge Assessment Development

In a parallel process, an 11-item knowledge assessment was developed with iterative feedback from subject matter experts. Rather than psychometric parameters assessing internal validity, knowledge assessments need to have external validity to ensure the most important concepts are being addressed. Core domains of knowledge gaps were identified from our previous qualitative work.(17) The questions were iteratively modified using informal cognitive interviewing and email feedback methods.

### Pilot Testing

The goal of the pilot testing was primarily to assess Usability and Acceptability of TRACH-Support across key stakeholders.(36–40) As such, no between group comparisons were made; rather, Pilot testing was conducted among providers (critical care physicians and advanced practice providers), critical care nurses, RTs, and surrogate decision-makers with both quantitative and qualitative collections. Please see **eTable 1** for details on recruitment for each participant group including inclusion/exclusion criteria. Surrogate decision-makers were recruited from a single academic safety-net hospital while providers, nurses, and RTs were recruited from multiple institutions via email invitations. Surrogates were approached to review and or use TRACH-Support if the patient was being considered for tracheostomy or had received MV for ≥7 days.

### Quantitative Data

REDCap, a HIPAA compliant online survey system, was used for all quantitative data collection.(41) Demographic data was collected for all participants. The co-primary outcomes for all participants were scores for the System Usability Scale (SUS) and the Acceptability of Intervention Measure (AIM).(42–46) Secondary outcomes included general impressions of TRACH-Support (all participants), Knowledge Assessment (surrogates), Intervention Appropriateness Measure (IAM) (providers, nurses, RTs), and Feasibility of Intervention Measure (FIM) (providers, nurses, RTs).(46) Participants with missing information were excluded. Preliminary efficacy data was also collected among surrogates including scores for the Decisional Conflict Scale (DCS).(47–54)

### Qualitative Data

A Think-Aloud approach was used for qualitative interviews in which participants were asked to verbalize their experiences around a specific task – in this case the use of TRACH-Support.(55–57) Participants were asked to focus on domains of Usability, Acceptability, and potential modifications (**eTable 2**). A matrix technique was used to analyze the data.(58–60) Interviews were conducted via a HIPAA compliant online conference platform by SL or ABM, audio recorded, and transcribed verbatim by a HIPAA compliant transcription service. An interview guide was created with a focus on Acceptability, Usability and recommended changes. Each page of TRACH-Support was reviewed with the participants with a focus on these concepts. A matrix focusing on these themes were created and Usability, Acceptability and any suggested changes was created. Each transcript had a primary reviewer who populated the matrix while a secondary reviewer assessed for any additions that were necessary. The goal was to recruit 10 surrogates and 10 providers for qualitative interviews.

The study was approved by the Colorado Multiple Institution Review Board (COMIRB 23-0171). All participants involved in pilot testing provided electronic consent for quantitative data collection and verbal consent for qualitative interviews. SAS v9.4 (Cary, NC) was used for quantitative data analysis while Atlas.ti V22 (Kreuzberg, Germany) was used for qualitative data management.

## Results

### TRACH-Support Development

The development phase was an iterative process engaging key stakeholders at multiple time points (**Figure 2**). The foundation of TRACH-support rested on extensive preliminary quantitative and qualitative work which identified several core informational gaps in the standard approach to decision-making (**Table 1**).(1,6,17,61) From the outset, TRACH-Support was designed to be used by multiple members of the multi-disciplinary team to guide surrogates through the decision-making process. Initially, a paper draft of the conversation tool was developed that mapped out the decision, risks/benefits, potential outcomes, and next steps. This paper draft was presented to several team members who made iterative modifications in the language and format. Subsequently, the paper draft was presented to a panel of shared decision-making experts (n=10). At this stage, there were suggestions around simplification of language, greater emphasis on the role of surrogate decision-makers, more in-depth representation of the options including a broader focus on time-trials, and representing potential outcomes in multiple ways. Based on this feedback, multiple changes were made and TRACH-Support was adapted to an online platform for greater accessibility. Additionally, a Navigation Pane (**Figure 3**) was introduced to increase customizability and allow users to move to different sections based on the flow of a conversation. The online version of TRACH-Support was presented to an existing panel of patients and surrogates that advised a patient centered decision-making working group. Feedback crossed multiple domains including appearance, appropriateness of language, readability, etc. A key comment was that the amount of information was overwhelming. It was suggested that information be presented based on the users needs. As such, TRACH-Support was iteratively modified to include pop-up windows where users could obtain more information about specific topics (e.g., different options, risks/benefits, etc.). Finally, TRAC-Support went through 2 additional revision phases including a group of critical experts and finally was re-presented to the existing panel of shared decision-making experts. It was during this phase that extensive outcome trajectories from a prior study was added.(1) This outcome data included numeric, verbal, pictorial, and time-lapsed outcome information for survival and death. The final version of TRACH-Support to be used in the pilot study (https://patientdecisionaid.org/trachsupport/introduction) was then evaluated based on IPDAS 74-item Criteria for Judging the Quality of Patient Decision Aids (**eFigure 1**).(62)

**Figure 3:**
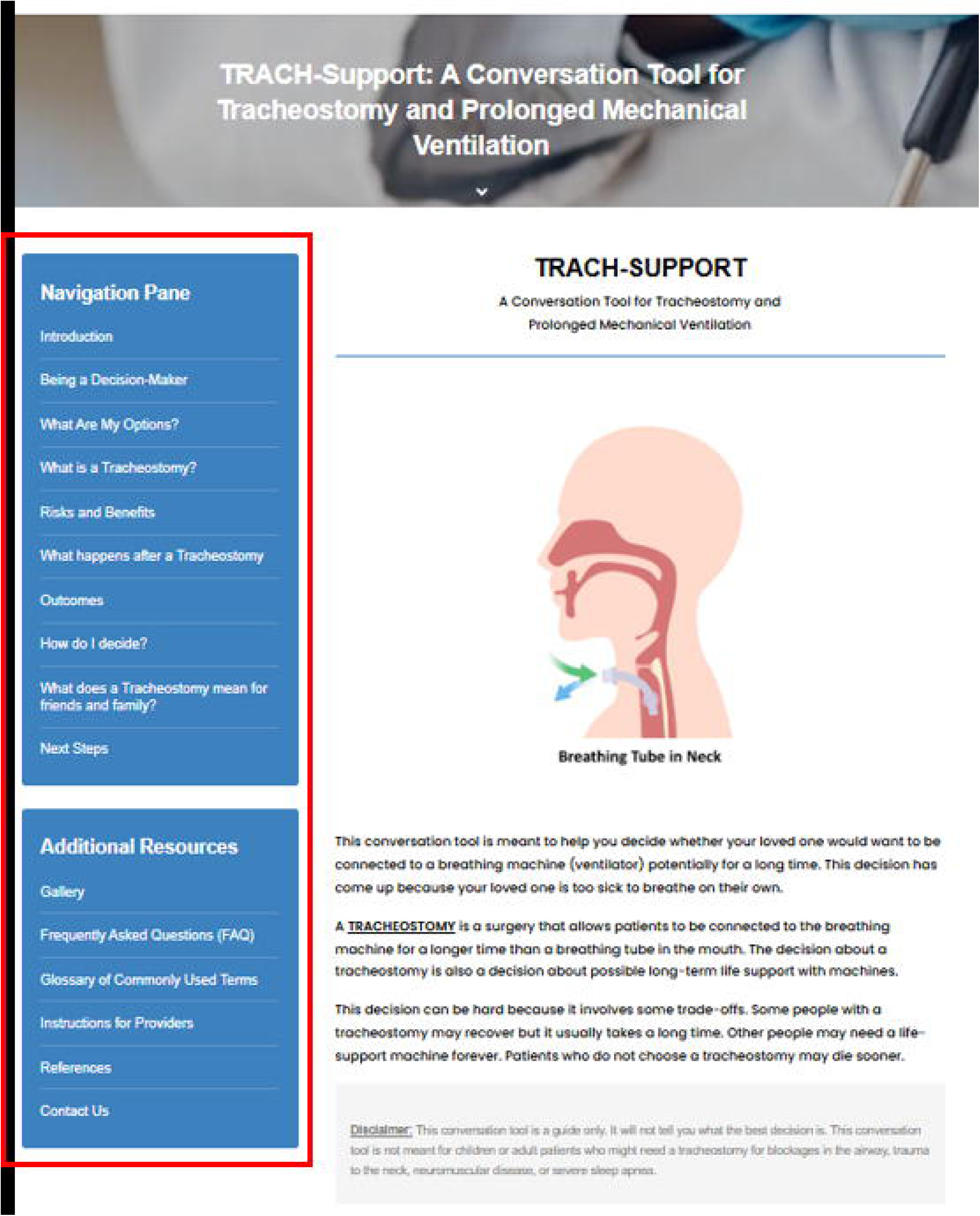
TRACH-Support Navigation Pane. – Figure 3 presents the introductory page for TRACH-Support. The Navigation Pane (red box) lists all the pages in TRACH-Support and is present on each page. It allows a user to go directly to the pieces of information needed most at a given moment in a conversation. This level of customizability allows each discussion about tracheostomy and prolonged mechanical ventilation to evolve on its own rather than a pre-arranged script like many past decision-support tools.

### Knowledge Assessment Development

Key surrogate knowledge gaps were identified based on prior work. The purpose of knowledge assessments is not to have internal psychometric validity but to have external validity at addressing the most important topics. A series of 13 true/false items were created based on these gaps and feedback was elicited from 10 subject matter experts via email responses or informal cognitive interviewing. Through this process, 2 items were dropped as they were viewed as confusing, redundant, or tangential to the primary purpose. The language for the remaining 11 items was revised to ensure literacy at a middle school level. The final version of the knowledge assessment was approved by the subject matter experts (**eTable 3**).

### Pilot Study

During the pilot study, a total of 86 individuals were recruited (15 surrogates, 29 providers, 31 nurses, and 11 RTs). Demographic data can be found in **eTable 4**. Across all participants the mean SUS=68.2/100 (SD=10.7) and the mean AIM=4.2/5 (SD=0.8). More than Among providers, nurses, and RTs the mean IAM=4.1/5 (SD=0.8) and mean FIM=4.2/5 (SD=0.6) (**Table 2**). 79.4% of participants viewed TRACH-Support as “Acceptable” or “Very Acceptable”, 74.2% viewed it as “Appropriate” or “Very Appropriate”, and 77.1% viewed it as “Feasible” or “Very Feasible”. All surrogates answered at least 9/11 knowledge items correctly (median=10/11, IQR=10-11).

**Table 2:** Pilot Study Outcomes.

|  | **Surrogate**<br>**(n=15)** | **Provider**<br>**(n=29)** | **Nurse (n=31)** | **RT (n=11)** | **Total (n=86)** |
| --- | --- | --- | --- | --- | --- |
| **SUS (mean**<br>**(SD))** | 74.2 (7.7) | 65.4 (10.3) | 69.2 (11.6) | 69.7 (9.7) | 68.2 (10.7) |
| **AIM (mean**<br>**(SD))** | 4.3 (0.4) | 4.1 (0.6) | 4.1 (1.1) | 4.4 (0.7) | 4.2 (0.8) |
| **IAM (mean**<br>**(SD))** | -- | 4.0 (0.6) | 4.1 (1.1) | 4.2 (0.6) | 4.1 (0.8) |
| **FIM (mean**<br>**(SD))** | -- | 4.2 (0.5) | 4.4 (0.6) | 4.1 (0.6) | 4.2 (0.6) |
| **Knowledge<sup>A</sup>**<br>**(median IQR)** | 10 (10-11) | -- | -- | -- | -- |

<sup>A</sup>11-item knowledge test. Number indicates the number of questions answered correctly by surrogates after reviewing TRACH-Support.

Participants were also asked about their general impressions about TRACH-Support (**Table 3**). Almost all participants had a positive experience with TRACH-Support (93.0% of “Positive” or “Very Positive”) and found it useful (97.7% “Useful” or “Very Useful”). Seventy-six (88.4%) participants would recommend TRACH-Supports to others. While most participants felt that the length and amount of information was “Just Right”, some providers, nurses, and RTs felt that TRACH-Support was “Too Long” and/or contained “Too Much” information. Among surrogates, the mean DCS=13.8 out of a possible 100.

**Table 3:** Participant Impressions of TRACH-Support.

| **Item (n (%))** | **Surrogate**<br>**(n=15)** | **Provider**<br>**(n=29)** | **Nurse**<br>**(n=31)** | **RT**<br>**(n=11)** | **Total**<br>**(n=86)** |
| --- | --- | --- | --- | --- | --- |
| **Very Positive or Positive Experience** | 15 (100) | 27 (93.1) | 28 (90.1) | 9 (81.8) | 80 (93.0) |
| **Very Useful or Useful** | 15 (100) | 28 (96.6) | 31 (100) | 10 (90.9) | 84 (97.7) |
| **Would You Recommend TRACH-Support?** |  |  |  |  |  |
| -Yes | 15 (100) | 22 (75.9) | 29 (93.5) | 10 (90.9) | 76 (88.4) |
| -No | 0 (0) | 0 (0) | 0 (0) | 0 (0) | 0 (0) |
| -I don't know | 0 (0) | 7 (24.1) | 2 (6.5) | 1 (9.1) | 10 (11.6) |
| **Length** |  |  |  |  |  |
| -Just Right | 15 (100) | 17 (58.6) | 26 (83.9) | 9 (81.8) | 67 (77.9) |
| -Too Long | 0 (0) | 12 (41.4) | 5 (16.1) | 2 (18.2) | 19 (22.1) |
| -Too Short | 0 (0) | 0 (0) | 0 (0) | 0 (0) | 0 (0) |
| **Amount of Information** |  |  |  |  |  |
| -Just Right | 15 (100) | 17 (58.6) | 26 (83.9) | 9 (81.8) | 67 (77.9) |
| -Too Much | 0 (0) | 11 (37.9) | 5 (16.1) | 1 (9.1) | 17 (19.8) |
| -Too Little | 0 (0) | 1 (3.4) | 0 (0) | 1 (9.1) | 2 (2.3) |
| **Balance** |  |  |  |  |  |
| -Balanced | 15 (100) | 21 (72.4) | 28 (90.3) | 10 (90.9) | 74 (86.0) |
| -Biased Towards Tracheostomy | 0 (0) | 2 (6.9) | 1 (3.2) | 0 (0) | 3 (3.5) |
| -Biased Against Tracheostomy | 0 (0) | 6 (20.7) | 2 (6.5) | 1 (9.1) | 9 (10.5) |

Qualitative interviews were also conducted among 10 surrogates and 10 providers using the Think-Aloud methodology with a matrix analytic approach. All participants interviewed indicated that they found TRACH-Support very useable and acceptable. The Navigation Pane that allowed for a customized approach to presenting information received high praise (*Provider 6: “I actually think this tool could be really great because we could start working through bits of the tool over, “Okay, so let’s just start introducing the idea of what this is that we’re talking about.” We could click on a few points and then be like, “All right. That’s a lot of information. We’ll talk more about it tomorrow and go through a few more points tomorrow.”)*. Additionally, participants also highlighted the pop-up windows for Options and Risks/Benefits as being a key method to access information on demand without having too much information one a single screen (*Surrogate 2: “I really liked the clickability”*). Several participants indicated that the outcomes data was highly useful (*Surrogate 3*: “*The rest of the information scanned it throughout the internet, it’s painful to hunt down when you’re actually looking at recovery times, the number of people who make it through, or anything like that. Yeah. This tool I like it a lot”).* Providers especially indicated that they liked the outcomes as it allowed them to feel more confident in their recommendations to families. Additionally, the numerous pictures were very helpful to both surrogates and providers. Many providers commented that despite having led multiple tracheostomy/PMV discussions, they had never shown a picture of a tracheostomy to surrogates.

Participants also identified gaps or had suggestions about additions that might improve TRACH-Support. Some had questions about long-term symptoms related to having a tube in the throat even after reviewing TRACH-Support (*Surrogate 2: “Does it hurt all the time? Is it numbing? Is that patient gonna be hurting all the time? Does the skin get irritated?”*). Some providers felt that being shown specific options based on values clarification early in the tool might be helpful (e.g., if comfort is the primary goal, then only showing comfort focused options). Both surrogates and providers felt that video testimonials would also be very impactful. Many providers suggested that certain sections were too wordy and that the tool would benefit from being shorter. Lastly, several participants recognized that faith or religion often influences the decision-making process and TRACH-Support does not address or acknowledge this fact.

## Discussion

Herein, we described the development and pilot testing of TRACH-Support, a novel web-based personalizable conversation tool for tracheostomy and PMV decision-making. We describe an iterative development process resulting in a decision-support tool with extensive key stakeholder engagement. Additionally, we present quantitative and qualitative pragmatic pilot study data that reveals high Usability and Acceptability for TRACH-Support.

The development phase for TRACH-Support adhered to best practice standards from the ODSF and IPDAS with engagement of patients, surrogates, critical care experts, and shared decision-making experts at multiple stages.(30–34,62,63) The broad engagement across multiple domains of subject matter experts and the inclusion of multiple different health care provider types expanded the development phase compared to previous decision-support tools in this space.(24,64) The web-based development also utilized best practices in User Centered Design with a heavy emphasis on graphics and pictures that likely contributed to the high Usability observed in pilot testing.(65–67) The iterative process ensured that stakeholders were able to make recommendations at each development stage.

There is significant controversy about how best to conduct pilot testing. The traditional view of preliminary efficacy and safety data in a small randomized controlled trial for a new medication is not feasible for behavioral or decision-support interventions especially when there are multiple targets for an intervention and when the intervention may be implemented across an entire unit. There is emerging consensus that pilot studies should focus on Usability and Acceptability and that traditional group by group comparisons may not always be possible.(36–40,68) As such, this pilot study was pragmatic in focusing on Usability and Acceptability across all anticipated users. The SUS score for TRACH-Support exceeded the average score for health apps previously published and the score among surrogates approached the category of “Excellence”.(69) Mean AIM, IAM, and FIM scores were also high with 8/10 participants viewing TRACH-Support as “Acceptable” or “Very Acceptable”. While no cutoffs exist for defining Acceptability thresholds, the high rating for TRACH-Support indicates that its design is highly successful.

From a usefulness perspective, almost all participants deemed TRACH-Support to be highly useful. This correlates with the fact that all surrogates answered at least 9/11 knowledge assessment questions correctly with more than half answering 10/11 correctly. These items were deemed to be critical information points needed during the decision-making process by panels of subject matter experts indicating a high level of information transference. Lastly, scores of the DCS are the most common primary outcome for larger clinical trials of decision-support tools.(50,51) The mean DCS in this study similar to if not better than the best rated end-of-life support tools, suggesting high preliminary efficacy.

The qualitative findings strongly support the quantitative findings. Key features that participants liked were the Navigation Pane customizability, clickability with pop-up windows, multiple graphics and pictures, inclusion of comfort care and time trial options, and the novel outcomes section. However, the qualitative data also revealed several items not evidence in the quantitative findings. While most indicated that the length was “Just Right” in the survey, many providers thought there were parts of the tool that were too wordy. Additionally, faith and religion as a factor in decision-making was noted to be missing by several participants. The addition of video testimonials was also felt to be a gap.

This study had several limitations. First, surrogates were only recruited from a single academic medical center which limits interpretations about broader Usability and Acceptability. Additionally, not all participants used TRACH-Support in a clinical encounter, some only reviewed it and applied it to prior decision-making scenarios. At the time of pilot testing, TRACH-Support was also only available in English. Therefore, its impact among non-English speaking surrogates is unknown.

In this pilot study, TRACH-Support was found to have high Usability and Acceptability among a broad representation of key stakeholders. Providers, nurses, and RTs viewed TRACH-Support as balanced, appropriate and feasible. Multiple additional content and design adaptions can be made to improve the overall experience. Future testing to determine effectiveness and implementation outcomes is needed and planned in a stepped-wedge cluster randomized trial.

## Supporting information

eTable

## Data Availability

All data produced in the present study are available upon reasonable request to the authors and in adherence with all regulations.

## Notes

### Competing Interest Statement

The authors have declared no competing interest.

### Funding Statement

This study was funded by the NIH/NHLBI K23HL141704

### Author Declarations

The Colorado Multiple Institutional Review Board of the University of Colorado gave ethical approval for this work.

