## Supplementary material for "Development and Pilot Testing for a Novel Shared Decision-Making Tool for Tracheostomy Decision-Making": eTable

**Online Supplement**

**eTable 1: Inclusion, Exclusion, and Recruitment Processes for Pilot Testing**

| **Participant Type** | **Site** | **Inclusion** | **Exclusion** |
| --- | --- | --- | --- |
| **Index Patient^A^** |  | Index Patient  -Age ≥ 18 years  -Acute or acute on chronic respiratory failure  -Tracheostomy already raised for patient OR  -MV ≥ 7 days | Exclusion  -Age < 18 years  -Pregnant  -Incarcerated  -Tracheostomy for progressive neuromuscular disease  -Tracheostomy for trauma |
| **Surrogate Decision-Maker^B^** | Single Academic Medical Center | -Age ≥ 18  -Surrogate identified by medical team  -English proficient | -Age < 18 years  -non-English proficient  -physician proxy  -court appointed guardian  -incarcerated |
| **Providers** | Multiple institutions recruited via email invitations | -Age ≥ 18 years  -Attending physician, fellow or advanced practice providers with specialty in critical care or palliative medicine | -Did not review TRACH-Support |
| **Nurses and Respiratory Therapists** | Multiple institutions via email invitations | -Age ≥ 18 years  -routine works in an critical care setting caring for patients receive mechanical ventilation  -in practice for ≥ 1 year | -Did no review TRACH-Support |

**eTable 2: Qualitative Interview Guide**

As with all Think-Aloud exercises, each page was reviewed with the participants with the following questions posed at various times. Feedback was elicited for each page.

**Qualitative Interview Guide**

Opening Script: Thank you for participating in this qualitative study to understand your experience with the tracheostomy decision-aid. This interview is being recorded. I want to make sure you know that there are no wrong answers. Anything you choose to tell us about your experience with the tracheostomy decision-aid will help us understand how to better help the decision-making process. All questions are optional and you can terminate the interview at any time if you feel excessive emotional distress. The interview will be less than 30 minutes. You have previously been provided information about consenting to being part of this study. Completing this interview is a sign of your informed consent. Do you have any questions prior to proceeding?

All Participants

1. Can you please tell me about your overall experience with the tracheostomy discussion(s)?
2. Can you please tell me about your experience with the tracheostomy decision-aid?
3. [Usability] Can you describe to me how useful the decision-aid was to you?
4. [Acceptability] Can you tell me about how easy the decision-aid was to use?
5. [Acceptability] Can you tell me what you thought about the language used in the decision-aid? Did you feel that the language was appropriate for you?
6. [Acceptability] Can you tell me about your thoughts on whether the decision-aid approached this emotionally complex decision appropriately?
7. Can you please tell me about what you learned from the decision-aid if anything?
8. Can you please tell me what aspects of the decision-aid were helpful?
9. Can you please tell me which aspects of the decision-aid were not helpful?
10. Are there any changes you would suggest for the decision-aid?

Providers Only

11. Can you describe to me how the tracheostomy decision-aid changed your usual approach to these types of conversations?
12. Have you ever used any other decision-support tools?
    a. IF YES – Describe to me how this decision-aid compared to other decision-support tools in the past?
13. Tell me your thoughts on using this decision-aid in the future?
    a. What might make you use or not use the decision-aid again?
14. Is there anything you want to tell me about your experience with the decision-aid?

**eFigure 1: IPDAS Patient Decision Aid Checklist**

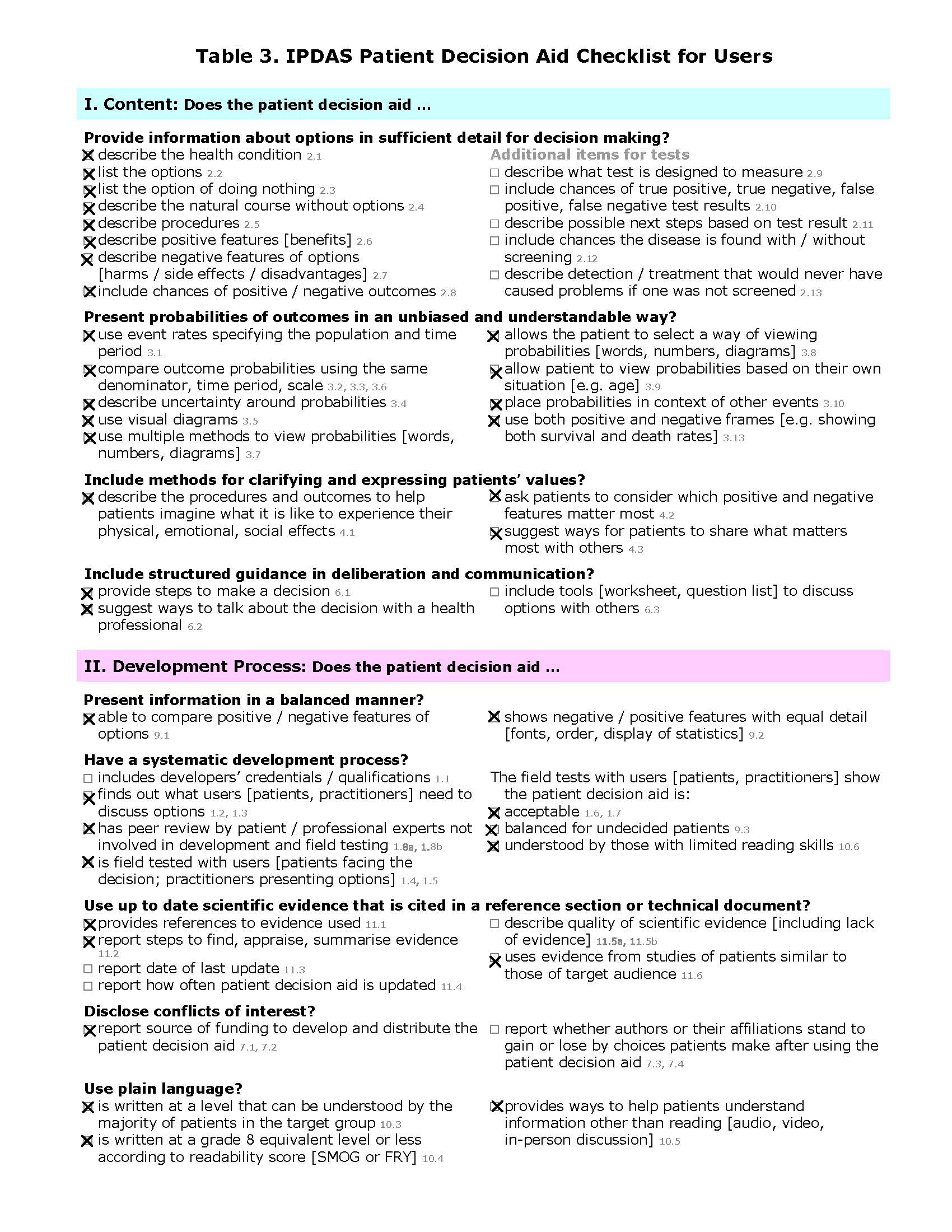

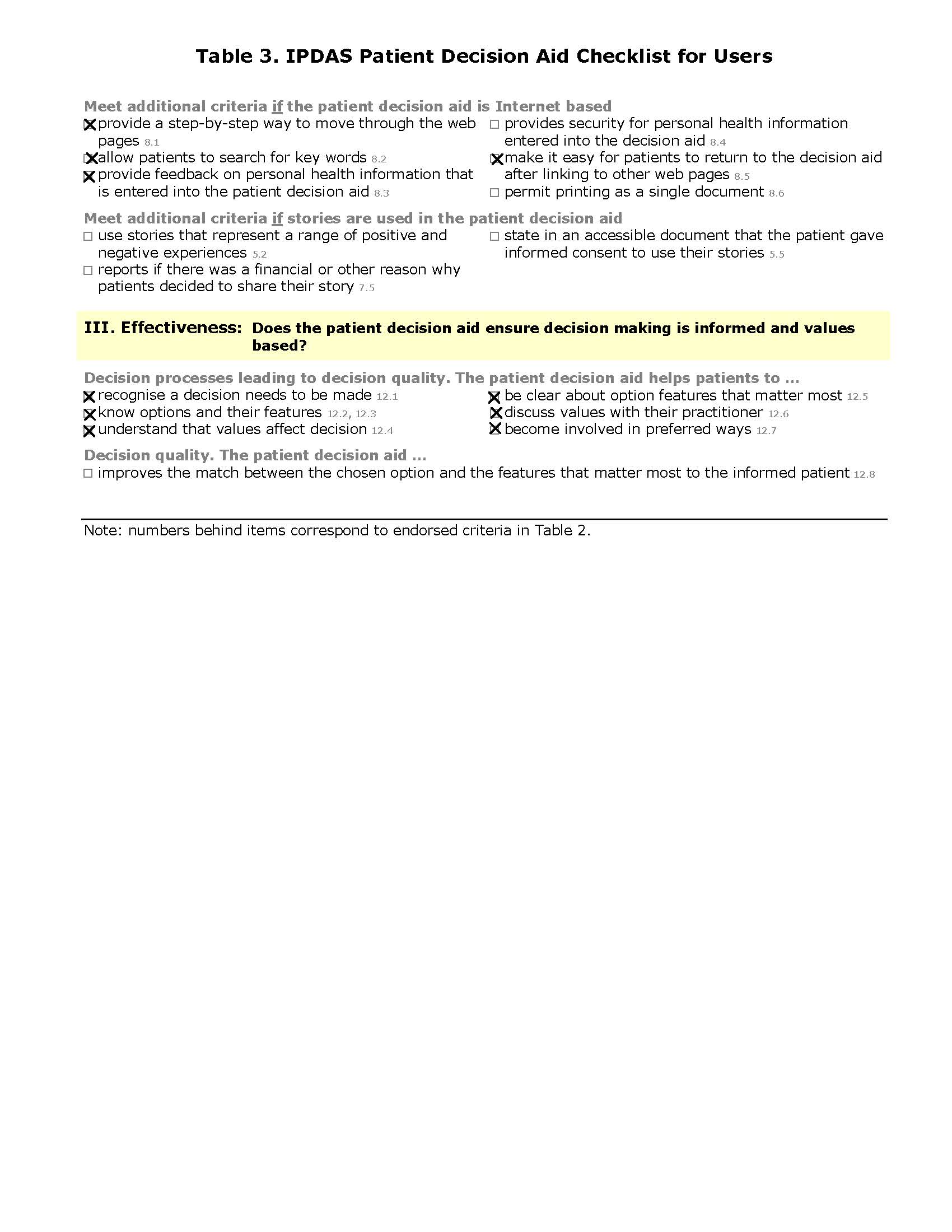

**eTable 3: Tracheostomy Knowledge Assessment for Surrogates**

|  | **Yes** | **No** | **Unsure** |
| --- | --- | --- | --- |
| 1. Tracheostomies help the lungs heal faster. |  |  |  |
| 2. Tracheostomies allow patients to be connected to breathing machines for a longer period of time. |  |  |  |
| 3. Breathing machines, or ventilators, are a form of life support. |  |  |  |
| 4. A tracheostomy is more comfortable for most patients than a breathing tube through the mouth |  |  |  |
| 5. Patients with tracheostomies are less likely to get infections in their lungs compared to patients with a breathing tube in their mouth |  |  |  |
| 6. After receiving a tracheostomy, it is easier for most patients to participate in physical therapy |  |  |  |
| 7. More than half of patients who receive a tracheostomy die within a year. |  |  |  |
| 8. Most patients with a tracheostomy are able to go home directly from the hospital |  |  |  |
| 9. Most patients with tracheostomies are able to go back to their previous normal function |  |  |  |
| 10. About half of patients under the age of 65 are able to return home 1 year after a tracheostomy |  |  |  |
| 11. Transitioning to a comfort focused approach is an alternative to tracheostomy. |  |  |  |

**eTable 4: Participant Demographics**

| **Item (n (%))** | **Surrogate (n=15)** | **Provider (n=29)** | **Nurse (n=31)** | **RT (n=11)** |
| --- | --- | --- | --- | --- |
| **Mean Age (SD)** | 53.6 (13.3) | 43.4 (8.3) | 40.6 (10.1) | 46.7 (12.0) |
| **Female, n (%)** | 13 (86.7) | 16 (55.2) | 23 (74.2) | 8 (72.7) |
| **Race, n (%)**  **-Asian**  **-Black/African American**  **-White**  **-Other**  **-Prefer Not to Answer** | 0 (0)  2 (13.3)  11 (73.3)  2 (13.3)  0 (0) | 2 (6.9)  0 (0)  26 (89.7)  0 (0)  1 (3.4) | 0 (0)  1 (3.2)  28 (90.3)  0 (0)  2 (6.5) | 0 (0)  2 (18.2)  9 (81.8)  0 (0)  0 (0) |
| **Years of Experience (%)**  **-<5 years**  **-6-10 years**  **-11-15 years**  **-16-20 years**  **->20 years** | -- | 7 (24.1)  8 (27.6)  6 (20.7)  4 (13.8)  4 (13.8) | 12 (38.7)  6 (19.4)  3 (9.7)  2 (6.5)  8 (25.8) | 1 (9.1)  1 (9.1)  3 (27.3)  1 (9.1)  5 (45.5) |
| **Prior Use of Decision-Support Tool? N (%)** | 1 (6.7) | 11 (37.9) | 0 (0) | 1 (9.1) |
